# Listening to mental health crisis needs at scale: using Natural Language Processing to understand and evaluate a mental health crisis text messaging service

**DOI:** 10.1101/2021.11.08.21266045

**Authors:** Zhaolu Liu, Robert L. Peach, Emma L. Lawrance, Ariele Noble, Mark A. Ungless, Mauricio Barahona

## Abstract

The current mental health crisis is a growing public health issue requiring a large-scale response that cannot be met with traditional services alone. Digital support tools are proliferating, yet most are not systematically evaluated, and we know little about their users and their needs. Shout is a free mental health text messaging service run by the charity Mental Health Innovations, which provides support for individuals in the UK experiencing mental or emotional distress and seeking help. Here we study a large data set of anonymised text message conversations and post-conversation surveys compiled through Shout. This data provides an opportunity to hear at scale from those experiencing distress; to better understand mental health needs for people not using traditional mental health services; and to evaluate the impact of a novel form of crisis support. We use natural language processing (NLP) to assess the adherence of volunteers to conversation techniques and formats, and to gain insight into demographic user groups and their behavioural expressions of distress. Our textual analyses achieve accurate classification of conversation stages (weighted accuracy = 88%), behaviours (1-hamming loss = 95%) and texter demographics (weighted accuracy = 96%), exemplifying how the application of NLP to frontline mental health data sets can aid with *post hoc* analysis and evaluation of quality of service provision in digital mental health services.

## 1 Introduction

Experiences of mental health difficulties and emotional distress are increasing globally and, according to the World Health Organisation, 700,000 people die by suicide world-wide each year. Even before the COVID-19 pandemic, the number of individuals meeting diagnostic criteria for emotional disorders or self-harming was growing in the UK, particularly among young women[1, 2]. The COVID-19 pandemic has only worsened this troubling picture with increased distress and mental health strain experienced by many individuals, and health systems struggling to meet demand[3]. Yet there is still much we do not know about the ‘who’, ‘what’ and ‘how’ of experiences of distress and mental health difficulties.

Evidence-based, good quality, timely and accessible support is needed to reach people in moments of distress and ensure they are heard, cared for, and equipped with appropriate resources. However, traditional mental health services, such as talking therapies provided through the UK National Health Service, cannot meet current demand, nor can they reach people in distress in a discreet and accessible format. To provide around-the-clock support, digital mental health services are increasingly being developed as an accessible alternative, or as adjunct resources to traditional face-to-face services. While such services hold great promise, and various digital mental health apps and tools continue to enter the market, few resources are properly evaluated, holding back progress in developing high quality support[4, 5].

The data from non-traditional digital mental health support services offers a unique window into mental health needs and experiences of distress, and provides an opportunity to evaluate personalised support approaches. In particular, big data analysis techniques, such as natural language processing (NLP) and machine learning, allow us to examine, at scale, expressions of distress and user interactions with mental health services[6]. To date, the challenges involved in accessing relevant data sets have partly prevented a full exploration of the value of such techniques for mental health insights and service evaluation (see [7] for a notable exception). The present work is an exception resulting from an in-depth collaboration between a research organisation (Imperial College London) and a frontline mental health service provider (Mental Health Innovations, MHI) with direct access to data, experts and volunteers.

In an effort to support individuals facing mental health difficulties, MHI launched Shout in 2019 as a 24/7 mental health crisis text line available to anyone in the United Kingdom (see mentalhealthinnovations.org/ impact-report-2021). The service connects highly trained volunteers with texters in distress. The text conversations aim to guide the texter to a calmer state and identify appropriate next steps. The volunteers are trained to follow a six-stage conversation structure that includes, in order: conversation initialisation (initialise); building rapport with the texter (build rapport); identifying the challenges that brought them to Shout (explore); identifying helpful next steps (identify goal); creating action plans (problem-solve); and closing the conversation (end the conversation). All conversations are overseen by clinically-trained supervisors, and over 900,000 conversations have been completed to date. Shout provides a discreet, anonymised, accessible and confidential mechanism to support individuals in distress, including those who are not in circumstances to express their feelings verbally but prefer texting for help. The textual data from each conversation is collected and undergoes an anonymisation process. Additional metadata and satisfaction ratings are collected from post-conversation surveys of volunteers and texters.

As the size of the Shout data set continues to grow, its value for understanding the mental health condition of the UK population increases. Additionally, there is a need to monitor and continually improve the quality of the service, while simultaneously increasing its scale as demand rises. Big data analysis methods offer the potential to understand at scale the texter population, their behaviour and experiences of the Shout service, and, in the long term, such models could potentially help monitor conversation progress in real-time.

Recent advances in NLP offer a valuable opportunity to interrogate the textual data collected by Shout[8, 9, 10, 11]. NLP is already present in our mobile phones and computers for tasks such as predictive text[12] or for translating between languages[13]. Recent NLP models learn an embedding (a numeric vector) from the textual data (words, sentences, paragraphs) which encodes contextual information. The quality of the output embedding depends on the language model, on the data used for its training, and the data to which it is applied (which should not be hugely dissimilar to the training data). The embedding can then be used for downstream tasks such as predicting expressed sentiment[14] or, in our case, predicting aspects about mental health conversations, such as texter demographics or behaviours.

NLP has been used in the context of medicine and healthcare, e.g., BioBERT[15] and Med-BERT[16]. In the context of mental health, NLP has also been used for predicting suicidal ideation[17, 18], analysing post-traumatic stress disorder[19], predicting psychosis[20] and other disorders[21], generating artificial mental health records[22], and for motivational interviewing[23]. For a more complete review of NLP in mental health, see [6]. However, NLP models have been been rarely applied to digital mental health resources such as crisis text line services[7], and to date no published studies analyse the Shout data set.

In this study, we use state-of-the-art NLP models to embed Shout conversation data followed by deep learning models to perform downstream tasks on these embeddings to better understand and evaluate the Shout service and its users. Specifically, we identified the following three focused tasks:

1. Predicting the **conversation stages** of messages.
2. Predicting the **behaviours** present in messages, from both texters and volunteers.
3. Classifying full conversations to extrapolate **demographic information** in texter surveys.

The first task helps us evaluate whether the Shout volunteers follow the conversation structure which they have been trained to follow. Success at this task would provide us with a tool that could be used in future for the analysis of which conversation structures or sub-stages are important for predicting outcomes, and how the structure of conversations differ across different texter or topic subgroups. The second task allows for exploration of the expressions of distress by texters (helpful for the understanding of mental health crises), and behaviours used by volunteers (helpful for ascertaining what behaviours are relevant for conversation outcomes) and could also be used to guide further research in future. The third task provides insights into the true user-base for Shout, controlling for the possible bias in survey completion across demographics. Understanding the true user demographic of Shout can help MHI to provide specific guidance to volunteers or, indeed, partner with other charities that can signpost appropriate resources more specifically target to particular subpopulations of users. It can also help identify which groups of texters Shout might not be reaching, allowing MHI to better target these underrepresented groups. In our study, we chose three demographic classes for prediction: ‘aged 13 and under’, ‘autism diagnosis’ and ‘non-binary gender’. These groups were suggested by MHI experts because they are either known to be higher risk for mental health challenges and may require tailored support or resources (for autism and non-binary gender), or for safe guarding (in the case of children 13 and under). Moreover, survey participation bias has previously been observed for gender [24, 25], age [26, 27] and disability [28, 29].

## 2 Materials and Methods

In this study, we aim to construct models that can learn the relationship between mental health conversation context (message text) and the labels of interest (such as survey outcomes or texter behaviour). In this section, we briefly introduce two essential components of our study: (1) the Shout data set and (2) the NLP models.

After providing some high-level statistics for the Shout data set, we describe the steps used to clean and filter the data set. We then describe the ground truth labels at the message and conversation levels that the NLP models will be trained to predict. Finally, we describe the necessary pre-processing steps for use of the conversation text within our NLP models.

For NLP and embedding of textual data, we use the Longformer model pre-trained on a large corpus of publicly available text. We detail the additional pre-training, where we updated the pre-trained weights using text from the Shout data set. We then describe the three experiments we performed in this study, where we fine-tuned the Longformer model to classify the chosen ground truth labels. We also describe hyper-parameter optimisation and our model evaluation measures.

### 2.1 Data

#### 2.1.1 Ethics

The study received ethical approval (19IC5511) from the Imperial College Research Ethics Committee (ICREC) and the data set was anonymised by MHI prior to researcher access. More explicitly, anonymisation removed any personally identifiable information such as names, phone numbers and addresses, by comparison with a curated word list. Moreover, researchers were trained in security and ethical considerations and were only able to access the data on a secure server.

#### 2.1.2 Shout service

The data was provided by Shout, a mental health crisis text line launched publicly by the digital mental health charity Mental Health Innovations in May 2019. Individuals of all ages who live in the UK can text 85258 to begin a conversation 24/7 whenever they are experiencing any mental or emotional distress and seeking help.

Once the conversation is initiated, the texter will receive an automatic reply from a bot asking for further information about the issues the texter is experiencing and stating the service terms and conditions. A trained Shout volunteer, who is supervised by a Clinical Supervisor, will be assigned the conversation and communicate with the texter following the initial messages. The volunteer will then respond to the texter and provide relevant support across six “conversation stages”, which they have been trained to follow in order (details of the stages can be found in Section 2.1.4). The conversation stages have been developed to help de-escalate a texter from a place of distress or overwhelm to a calmer place from where they can move forward.

After the conversation, texters are sent a link to an optional post-conversation survey that contains various questions about the texter’s experience of the conversation; what brought them to Shout; and their demographic information. In our data sample, 13.75% of texters completed the survey on at least one of their conversations with Shout. The volunteer also completes a short survey after every conversation providing information about the key topic categories raised in the conversation, and whether the texter experienced and expressed any level of suicide risk.

#### 2.1.3 Conversation textual data

Our analysis was conducted on data recorded from 12 February 2018 to 3 April 2020, comprising 271,445 conversations and 10,809,178 messages. Conversations were of varying length (Figure 1A). In very short conversations, it was assumed that the texter did not engage with the conversation, as the conversation would be largely comprised of standard system messages from the automated bot. Very long conversations are generally rare and atypical: long conversations typically appear under very high risk scenarios, and, when such risk is not present, are discouraged due to the limited availability of volunteers and the importance of closing the conversation so the texter can move forward with any agreed next steps.

**Figure 1:**
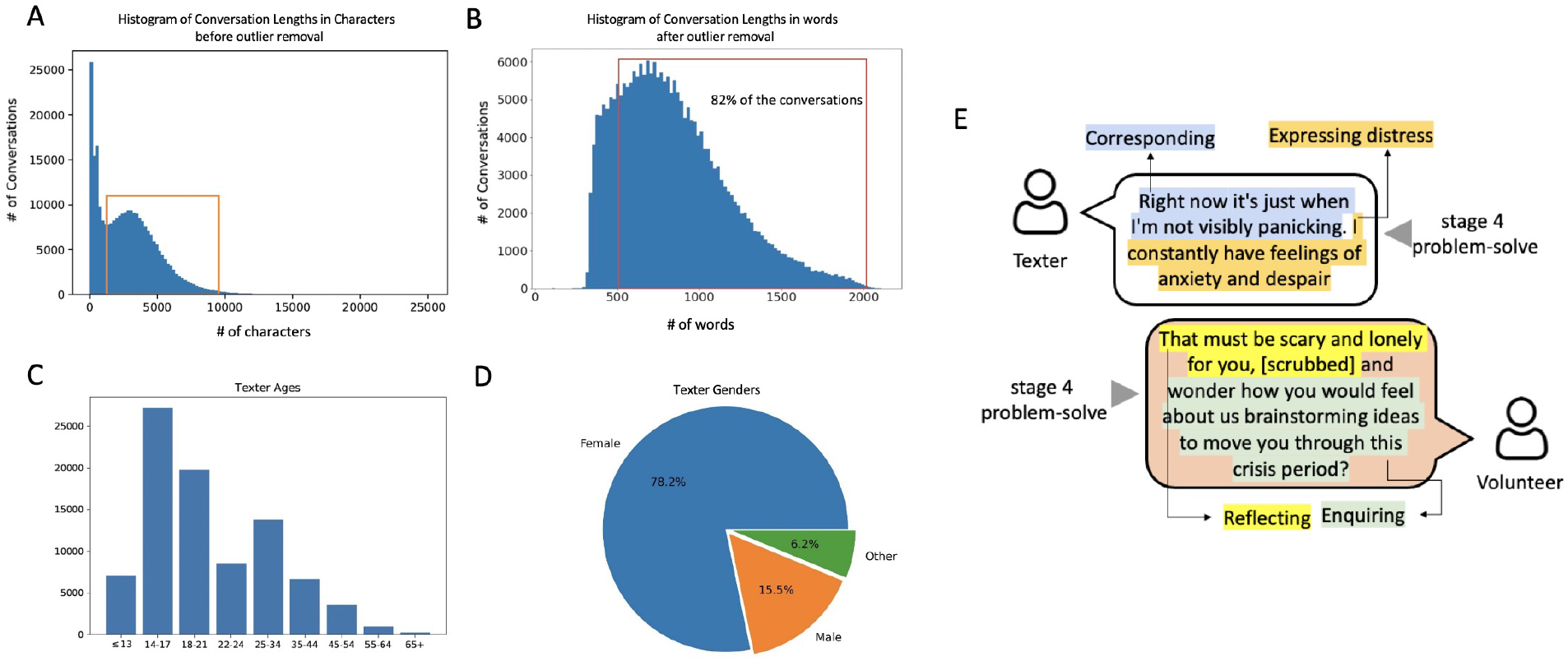
High-level overview of the Shout data set. **A** The distribution of conversation lengths as measured by the number of characters. The red box indicates the conversations between 1200 and 9200 characters which passed the inclusion criteria for analysis. B The histogram of the number of words in a conversation. Since 82% of the conversations exceed the maximum word limit (512 words) of traditional BERT-style NLP models, we use the Longformer model. C Age distribution and D gender distribution of texters as obtained from the texter surveys (which we show in Section 3.3 to be biased towards particular demographic groups). E A synthetic example of a portion of a conversation to illustrate annotations of behaviours and conversation stages.

In addition to the raw textual information, recorded meta-data includes the time-stamp, the actors (texter, volunteer or Clinical Supervisor), and Volunteer ID. To provide the reader with an intuition for the Shout conversations, we provide a short constructed example *emulating* a Shout conversation in Figure 1E.

##### Filtering the conversations

Conversations were filtered to remove extremely short (texter did not engage) or extremely long conversations (high-risk scenarios); hence the peaks to the left of the red box and the tail to the right of the red box in Figure 1A were treated as outliers and removed (85,803 conversations). The resulting data set used for analyses in this paper is a sub-set (68.4%) of the total conversations which fell within the range of 81-2129 words (as shown in Figure 1B).

#### 2.1.4 Ground truth labels

We use supervised learning NLP models in this study, for which it is necessary to train and test the model against ground truth data. To train a model to predict conversation stages and texter/Volunteer behaviours (tasks 1 and 2), we use annotations at the individual message level, performed by expert Clinical Supervisor and research psychologist (AN). To train a model to predict texter demographics from the entire conversation content (task 3), we use as ground truth labels the texter survey responses available.

##### Message-level labels

A subset of 169 conversations (8844 messages) was randomly selected from the 271,445 total conversations and annotated at the message level to indicate the corresponding ‘conversation stage’. The annotation was performed by the Shout Clinical Supervisor and research psychologist, according to the definitions of each conversation stage established by Shout to train Volunteers. Each message belongs to one of six conversation stages listed in Table 1, with stage 0 incorporating preliminary engagement of the texter with the Shout service and automated bot, and stages one to five comprising the conversation with the Volunteer.

**Table 1:** Pre-defined conversation stages which Shout Volunteers are trained to follow to best support the texters

| Stage code | Name | Brief definition |
| --- | --- | --- |
| 0 | Initialise | Greetings and bot messages |
| 1 | Build rapport | Active listening and good contact techniques. |
| 2 | Explore | Understand what is the crisis and asses risk. |
| 3 | Identify goal | Clarify what support the texter needs. |
| 4 | Problem-solve | Identify current resources and create an action plan. |
| 5 | End the conversation | Review the action plan and close warmly |

Similarly, the Shout clinical supervisor and research psychologist (AN) annotated the behaviours present in each message. A process of codebook development[30, 31] was applied in an inductive thematic analysis[32, 33]. Codebook development is a well established methodology in qualitative research, which allows for iterative strengthening of the definitions through interactions with other members of the team, and leads to a structured trail of evidence and enhanced replication of findings. Specifically, once working definitions were developed for each theme, two other MHI Clinical staff applied the codebook to the data. These additional Shout staff independently coded a subset of the data as a validity check [34, 35], and in the case of any initial coding discrepancies the definitions associated with relevant codes were updated and refined. Therefore the codebook was iterated until there was convergence in coding agreement between the Shout researchers. As a result of this process of iteration, a total of six distinct behaviours were identified and defined, namely: setting intention, enquiring, expressing distress, reflecting, corresponding, and discord.

Messages in the 169 conversations comprising our sample were annotated with up up to a maximum of 3 behaviours for each message, in the order in which they appear. We provide further definitions of the behaviours in Table 2.

**Table 2:** Definitions of behaviours

| Behaviour code | Name | Brief definition |
| --- | --- | --- |
| 1 | Setting intention | Communicate an immediate (near future) aim |
| 2 | Enquiring | Explore one’s understanding of another’s experience |
| 3 | Expressing distress | Communicate an offloading of negative feelings |
| 4 | Reflecting | Mirror something the texter has said |
| 5 | Corresponding | Show comparability between both parties |
| 6 | Discord | Involves a lack of harmony between both parties |

##### Conversation-level labels

After the conversation, texters have the option to complete a survey that contains questions about the texter’s demographic information (including age, gender identify, ethnicity and disability), and several questions related to the conversation such as “Did you find the conversation helpfulã”. In our data set, only 13.75% of the conversations had an associated completed texter survey, meaning that there is large uncertainty as to the demographic profile of the texters. In addtion, previous studies suggest there may be completion bias in texter surveys[36, 37]; therefore it is unreliable to extrapolate the distribution of demographics from the texter survey to the entire data set. Figure 1 (C) (D) shows the distributions of age and gender in the texter survey. We used the demographic labels in the available texter surveys to train a model to map conversation content to demographic labels.

Here, we focussed on three classification categories, namely 1) self-declared autism diagnosis; 2) self-declared nonbinary gender; 3) aged 13 and under. These were chosen to both understand mental health needs in the population, and help Shout identify use of their service by groups for whom more tailored support and resources may be warranted.

#### 2.1.5 Pre-processing

##### Special tokens

The Shout data set is a collection of conversations that involve three ‘actors’: the texters, the volunteers and the Shout bot (automated messages generated for initiating and ending conversations). The tokens “[texter]”, “[Volunteer]” and “[bot]” are added in front of the messages from the corresponding speaker to separate the text of the different actors.

##### Data augmentation for messages

Due to the small sample size of annotated messages (169 conversations comprised of 8844 messages), data augmentation techniques (e.g., word deletion, word swapping and synonym replacement) were employed to boost performance and ensure robustness of the trained model[38]. The augmented messages retain the same label as the original messages, as it has been empirically shown that augmented data obtained from simple deletion, swapping, and replacement inherit the label from the original labeled text [38].

### 2.2 Computational methods

#### 2.2.1 NLP: Longformer

The Longformer NLP architecture was used to model the conversation content[39] since 82% of the pre-processed conversations exceeded the 512 word limit of standard NLP models [10, 11]. The Longformer model uses specially designed attention patterns (local attention and global attention) to cope with long word sequences. The Longformer model was implemented via Python using the Huggingface package[40], which comes with pre-trained weights[39], i.e., the model had already been trained with a large corpus of text comprised of a wide selection of non-domain specific documents found across the internet.

#### 2.2.2 Additional pre-training

In addition to the pre-training performed by Huggingface, the Longformer checkpoint[39] was further pre-trained on the masked language model (MLM) using the entire pre-processed Shout data set (see Figure 2). Words within a text sample are randomly masked; predicted from their surrounding context; and predictions evaluated against the actual masked word. This additional pre-training updates the weights to account for deviations of our data set from the original, non-specific text corpus used by Huggingface.

**Figure 2:**
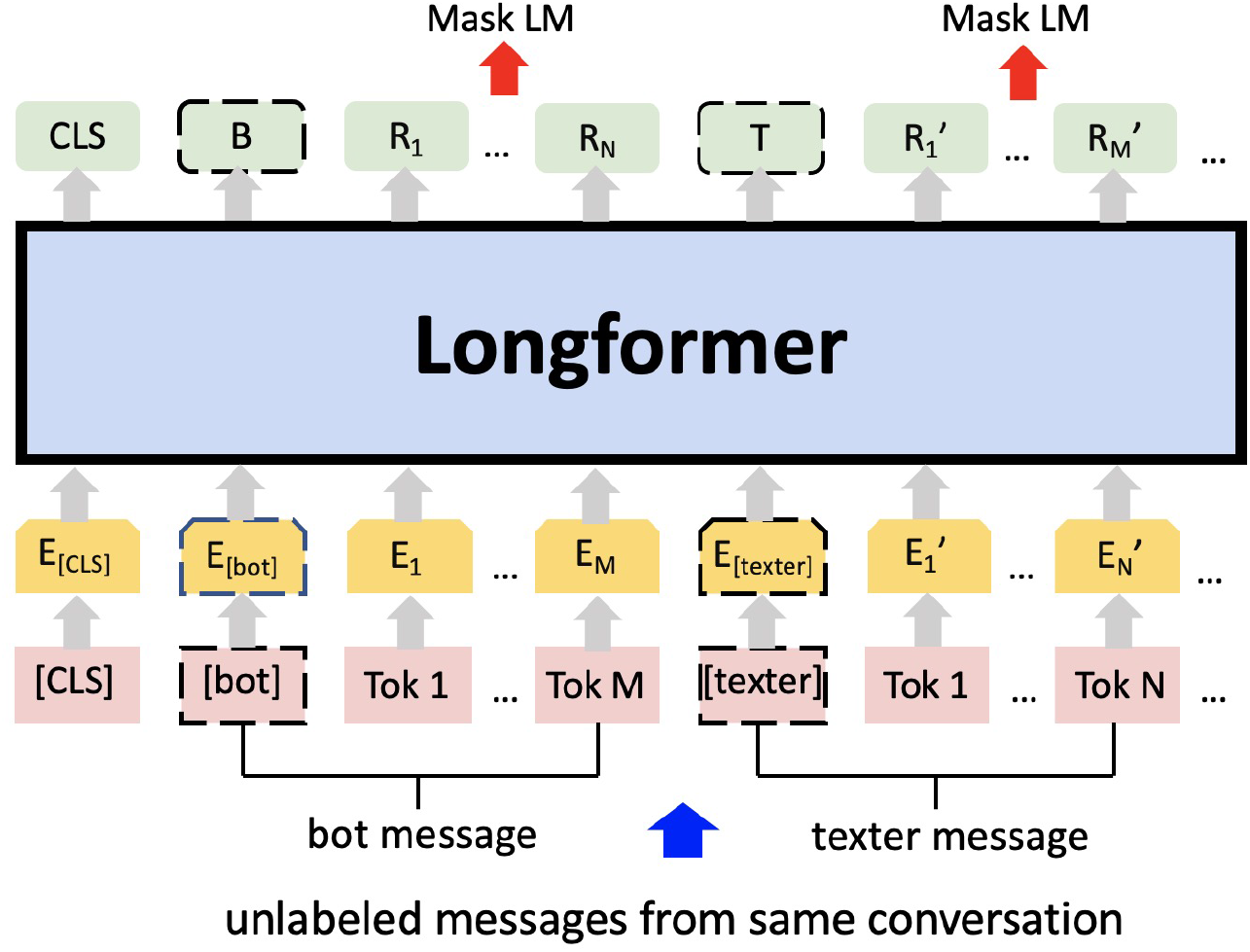
Additional pre-training on the masked language modelling (MLM) task using Shout mental health data. *E* represents the input embeddings. *R*_*i*_ represents the contextual representation of token *i. B* and *T* are the contextual representations of special token [bot] and [texter] respectively. After the tokenisation the numerical vectors are processed via Longformer.

The maximum conversation length is set to 2048, as 99.98% of the conversations have less than 2048 words. Padding and truncation were done for conversations less than 512 words or greater than 2048 words, respectively.

#### 2.2.3 Fine-tuning and classification

After pre-training, the Longformer model was trained for classification of the chosen labels. In order to inherit the learned weights from the MLM after the additional pre-training phase described in Section 2.2.2, the language modeling head (i.e., the top layers for generating the predictions of masked words) is removed while the other layers and their corresponding weights are retained. In other words, we keep the encoder that generates the numerical embeddings of the text, and remove the decoder that converts the numerical embeddings back into text. An additional classification layer with a non-linear activation function (Sigmoid or Softmax) was then added to the encoder. This additional layer provides a set of additional parameters that can be learnt to map the numerical embeddings (encoded text) to the ground truth class labels. The choice of non-linear activation function for the additional classification layer depends on the task; for example, multi-class classification requires a Softmax activation function which normalizes the output into a probability distribution.

We carried out three experiments aimed to classify three different labels to match the identified tasks outlined above:

##### Experiment 1: classification of conversation stages

This is a message-level multi-class classification task, where one of the six conversation stages (Table 1) is assigned to a given message. In this experiment, the non-linear activation function for the additional layer was a Softmax function.

##### Experiment 2: classification of behaviours

Up to three different behaviours were assigned to each message, reflecting the fact that texters and volunteers may display multiple behaviours within a text message. Hence this is a message-level multi-label classification task, where the six behaviours (Table 2) are represented by six independent binary labels. To generate independent probabilities for each label, the non-linear activation function used in the classification layer was a Sigmoid function. Note that in this task of classification of behaviours, data augmentation was not used, as perturbations to the messages can change the multi-label structure.

##### Experiment 3: classification of texter demographics

Texter demographics were recorded in the texter survey at the conversation level. We chose categories recorded in the Shout data set in line with demographics of particular interest to MHI. The binary demographic categories (model labels) are: age (13 and under or over 13); autism (does the texter self-identify as having an autism diagnosis or not); and non-binary gender (does the texter self-identify as either agender, genderqueer, trans female, or trans male). In a similar manner to Experiment 1, the top layers of the Longformer model were substituted by a classification layer followed by Softmax activation function.

#### 2.2.4 Hyperparameter optimisation

In addition to learning the parameters of the Longformer model, there are various hyper-parameters that must be pre-defined. Here, we describe the choices of hyper-parameters and optimisation procedures for the pre-training and for the learnt classification models.

##### Hyper-parameters of MLM pre-training

A learning rate of 5 × 10^−5^ was chosen as it has been proved successful in previous relevant studies[10, 39, 40]. The number of epochs (3) and batch sizes (2) were adopted as a reasonable choice given the computational power available.

##### Hyper-parameters of classification models

The hyperparameters for the three classification models (Experiments 1-3) were optimised through a grid search of learning rate ([8 · 10^−6^, 10^−5^, 2 · 10^−5^, 5 · 10^−5^]), number of epochs ([3, 4, 5]), and batch size ([2, 4, 8, 16]). The parameter combination yielding the best classification performance for all three classification models was: learning rate= 8 *·* 10^−6^, number of epochs= 5, batch size= 8.

#### 2.2.5 Evaluation and Performance

To evaluate the quality of the classification models, we use weighted accuracy, i.e., accuracy averaged for each class[41]. For the behaviour classification model, we also report two additional measures. The (1 - Hamming loss), where values closer to 1 imply better results[42], provides a more meaningful output for multi-label classification as it considers the fraction of labels that are incorrectly predicted, whereas standard weighted accuracy would require all labels for a particular example to be correct to be considered accurate. The label ranking average precision score (LRAP) is a multi-label ranking metric that gives higher scores to predictions closer to the ground truth labels[43].

## 3 Results

### 3.1 Classifying messages into conversation stages (Experiment 1)

We first evaluated our ability to classify messages into the six conversation stages (Table 1), using a multi-class classification model. Here, a message could only be assigned to a single conversation stage label.

#### 3.1.1 Model optimisation and prediction performance

The weights from the MLM were fine-tuned (see Section 2.2.3) against the conversation stage class labels. For robustness, and to provide insights into the necessary components for learning mental health conversations, we examined the effects of changing: (i) the percentage of data used to pre-train the MLM; (ii) the percentage of training data used to fine-tune the model for conversation stage classification; (iii) the use of text augmentation; and (iv) the inclusion of text from both the preceding and subsequent messages to aid classification (see Figure 3 for a visual description of including additional context).

**Figure 3:**
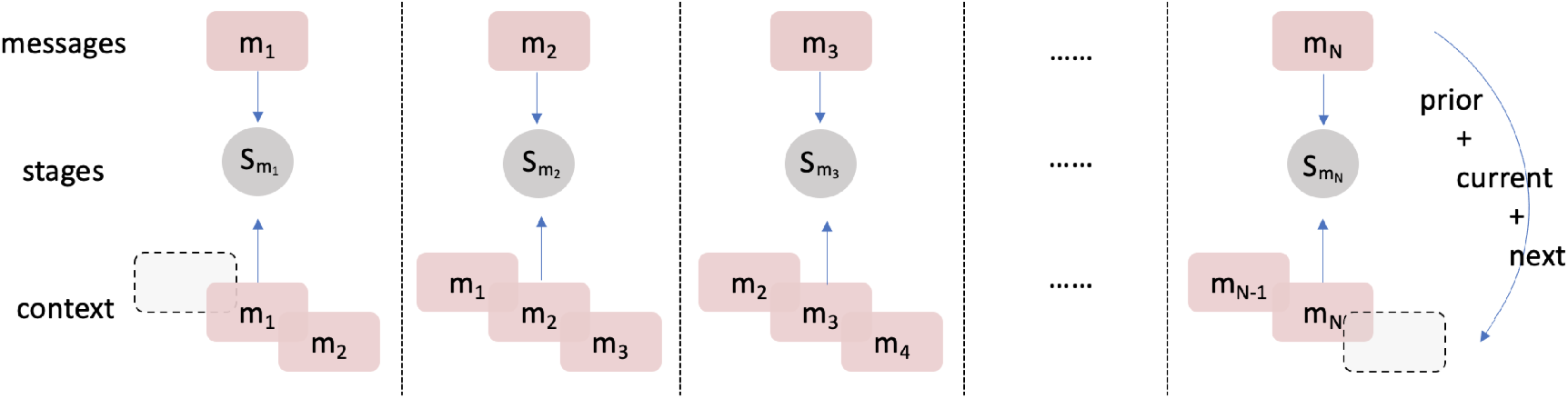
Mechanism to include context in input data: *m*_*i*_ represents the message at time *i* and *S*_*j*_ represents the stages assigned for message j. Exactly one prior and one post message are concatenated with the message of interest. The label of the contextualised text inherits the original label of the current message.

Table 3 shows our results for the different models trained. The optimal model (model 3) achieved accuracy of 87.75%, and was pre-trained on 100% of textual data, used 100% of training data for fine-tuning, included text augmentation, and included context of the preceding and following messages. The different classification performance of the seven models in Table 3 allows us to understand the importance each component. The hyper-parameters of all models were optimised using 5-fold cross validation, see Section 2.2.4.

**Table 3:** Comparison of conversation stage classification performance for models with different performance improvement techniques. ‘No context’ means that only the message of interest is input to the model, whereas ‘± 1 message’ refers to the context concatenation scheme introduced in Figure 3, wherein we included the text from the messages preceding and subsequent to the message of interest.

| Model ID | % Pre-training data | % Classification training data | Augmentation | Context | Accuracy |
| --- | --- | --- | --- | --- | --- |
| 1 | 100% | 100% | × | No context | 0.6231 |
| 2 | 100% | 100% | × | ± 1 message | 0.8620 |
| **3** | 100% | 100% | ✓ | ± 1 message | **0.8775** |
| 4 | 0% | 100% | × | ± 1 message | 0.2010 |
| 5 | 33% | 100% | × | ± 1 message | 0.7512 |
| 6 | 100% | 25% | × | ± 1 message | 0.6364 |
| 7 | 100% | 50% | × | ± 1 message | 0.6966 |

Table 3: Comparison of conversation stage classification performance for models with different performance improvement techniques. ‘No context’ means that only the message of interest is input to the model, whereas ‘± 1 message’ refers to the context concatenation scheme introduced in Figure 3, wherein we included the text from the messages preceding and subsequent to the message of interest.

From the trained models in Table 3, we draw four main observations. First, we find that without pre-training, the accuracy of the MLM (model 4, acc=0.2010) is significantly lower than pre-training with just 33% (model 5, acc=0.7512) or 100% (model 2, acc=0.8620) of the total avaliable textual data. This result confirms the importance of conducting additional pre-training on the raw conversation text to improve the model’s understanding of text specific to the Shout data set.

Second, increasing the proportion of training data for fine-tuning the model for classification improves accuracy (model 6, acc=0.6364 *<* model 7, acc=0.6966 *<* model 2, acc=0.8620). This result is expected as training with more data typically results in higher accuracy. However, models trained with substantially smaller data sets (models 6 and 7) also produced reasonably accurate outcomes.

Third, using text augmentation increases the accuracy of message classification (model 2, acc=0.8620 *<* model 3, acc=0.8775). Data augmentation is used in most implementations of deep learning algorithms, most commonly as a regularisation strategy to prevent overfitting[44]. In NLP, learning of high-frequency numeric patterns (e.g., token embeddings) or memorisation of particular forms of language prevent generalisation. Therefore, we would expect an increase in accuracy with augmentation. Since texters often use very informal language via their phones augmentation reduces the likelihood of the model overfitting to the unique writing styles present in a subset of conversations that may not generalise to a test set of conversations.

Fourth, using additional context to classify the message of interest improves accuracy (model 1, acc=0.6231 *<* model 2, acc=0.8620). Clearly, the model stands to benefit from the extra context surrounding the target message because changes in conversation stage are relatively slow (i.e. multiple consecutive messages in a short time period are likely to have the same conversation stage label). The input data ‘± 1 message’ scheme therefore includes three times the information of the original target message for the conversation stage, resulting in better performance.

#### 3.1.2 Classification performance of individual conversation stages

Given that we achieved high classification accuracy for conversation stages, we now ask whether the classification accuracy of individual stages differed. In Table 4 we show the classification accuracy for each conversation stage. We find that the classification accuracy for each stage is comparable to that of the entire model, and that the variability among conversation stages is minimal (std = 0.009), suggesting that our model has generalised to all conversation stages.

**Table 4:** Classification accuracy for each conversation stage with the optimal model (Model ID 3 in Table 3).

| Stage Name | Initialise | Build rapport | Explore | Identify Goal | Problem Solve | End |
| --- | --- | --- | --- | --- | --- | --- |
| **Accuracy** | 0.9001 | 0.8890 | 0.8936 | 0.8995 | 0.8709 | 0.8929 |

#### 3.1.3 Feature analysis and interpretation

Despite boasting high accuracy predictions, deep learning methods often do so at the expense of interpretability. To get insight into mental health needs and factors affecting Shout service quality, it would be highly valuable not only to predict labels of interest, but also to understand what features of the conversations and language used are linked to these labels. To generate such insights, we used LIME (Local Interpretable Model-Agnostic Explanations) to provide ‘explanations’ of the decisions of the trained model [45]. LIME learns local information required to make a prediction and ranks individual words according to their contribution. We used LIME to extract the two most important words contributing to the final prediction of 300 random samples for each conversation stage (total of 600 words per conversation stage). Plotting word clouds of these word collections allows for a visual interrogation of which words the model finds important for predicting the conversation stage label. Below we provide summaries of the distinct features of each conversation stage according to the word clouds (Figure 4a-f for conversation stages 0-5):

**Figure 4:**
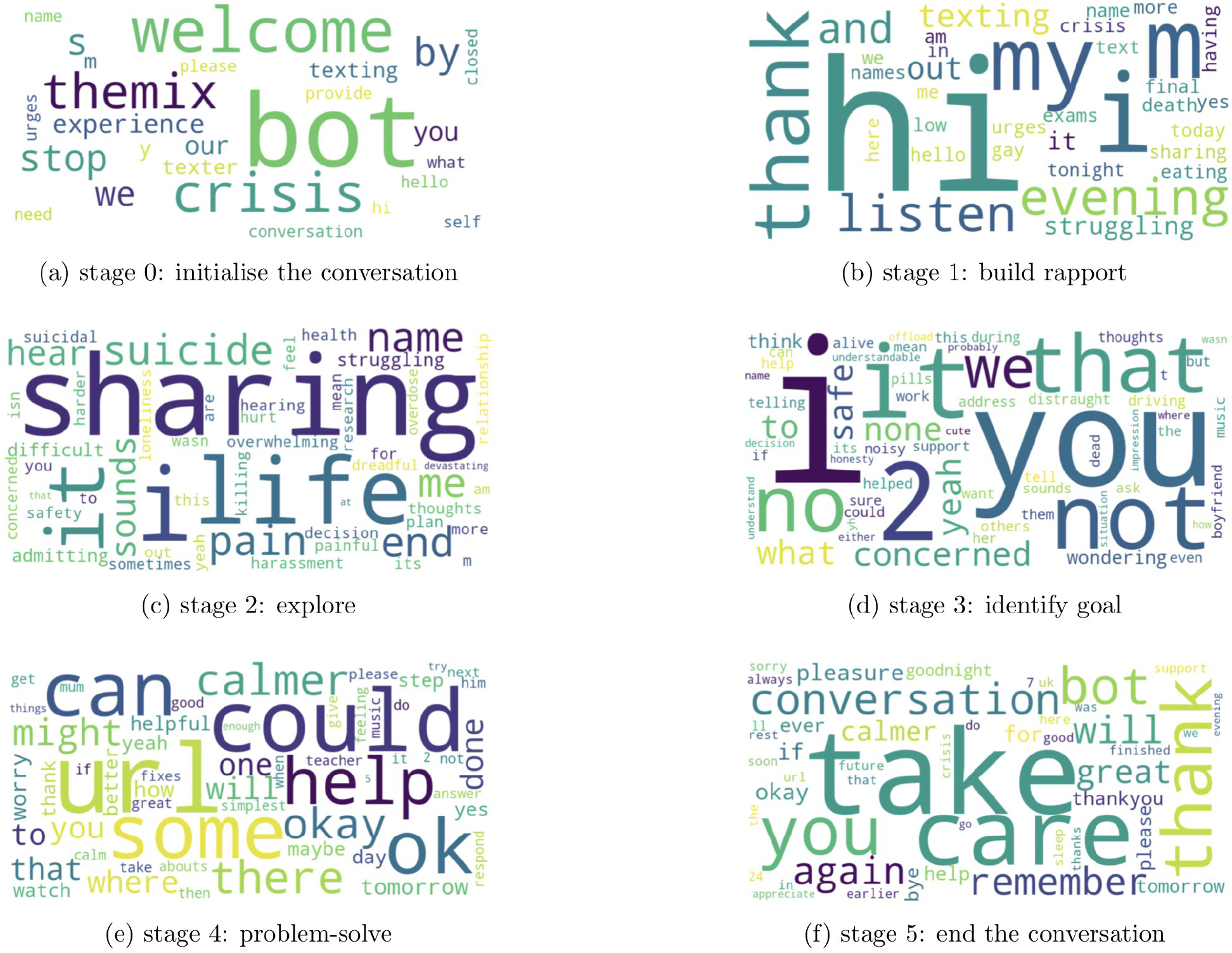
Word clouds for LIME-derived words that best explain the model prediction for each conversation stage. **(a)** Bot messages are detected from the *initialisation*. **(b)** Greeting words are frequent when *building rapport*. **(c)** Volunteer begins to *explore* the texter’s problem with key words from the discussion featuring ‘sharing’ and ‘life’. **(d)** Volunteer works to *identify goals* with the texter and these messages usually start with first-person and second-person pronouns.

- Stage 0 - *Initialisation*: Dominated by introductory words (‘welcome’ and ‘hello’) and includes messages from the ‘bot’ and the ‘texter’, showing that the model has learnt that Stage 0 is associated with the beginning/initialisation of the conversation.
- Stage 1 - *Build rapport*: Includes introductory words such as ‘hi’ and ‘evening’, but this time from the volunteer rather than the texter. Stage 1 also includes words that begin to explore how the texter is feeling as can be seen from ‘struggling’ and ‘crisis’.
- Stage 2 - *Explore*: Volunteers are trying to explore the problems in detail; we can see words such as ‘sharing’ and ‘life’, alongside descriptive terms such as ‘pain’, ‘suicide’, ‘overwhelming’, ‘difficult’ and ‘killing’.
- Stage 3 - *Identify goal*: After establishing the problem, Volunteers try to clarify what support the texter is seeking and these messages usually start with first-person and second-person pronouns such as ‘I’, ‘we’ and ‘you.’ The goals identified at this stage appear to be texter-specific and no recurrent adjectives and nouns are present in the word cloud.
- Stage 4 - *Problem-solve*: Volunteers explore with the texter what they can identify as helpful steps to take after the conversation, from small things like listening to music, to bigger things like talking to a trusted person about their experiences and difficulties. Volunteers may also signpost texters to relevant further resources or organisations to provide ongoing support or information. Suggestions generally include words like ‘can’, ‘could’ and ‘might.’ ‘Help(ful)’ resources can include signposts with a ‘URL’.
- Stage 5 - *End the conversation*: Volunteers will bring the conversation to a close with phrases like ‘take care’, ‘pleasure’ and ‘bye’. Texters express gratitude by saying ‘thank you’ and ‘thanks.’

Hence the word clouds are closely related to each conversation stage and the consistency of LIME-derived features for each class label means that the accurate predictions made by the model are based on meaningful and interpretable features of the messages, in line with similar features used by the human annotator when assigning class labels. **(e)** Texter-specific *solutions* are suggested such as a ‘helpful’ ‘URL’. **(f)** The conversations are brought to a close using phrases like ‘take care’ and ‘bye’.

### 3.2 Predicting texter behaviour expressed in messages (Experiment 2)

In the previous section we showed that the conversation stage for a message could be accurately classified using a multiclass model architecture. In this section we explore the classification of messages into different psychologically-relevant ‘behaviours’, as discussed in Section 2.1.4 and defined in Table 2. In contrast to Experiment 1, where messages could only be assigned to a single conversation stage, multiple behaviours can be present in a single message and therefore can be annotated with multiple (up to a maximum of three) behaviours. Therefore, we used a multi-label classification architecture, where the six behaviours (Table 2) are represented by six independent binary labels.

#### 3.2.1 Model optimisation and prediction performance

Comparing models with and without message context (Table 5), we find that the optimal model does not use the text from the preceding and subsequent messages: all three metrics for the ‘no context’ model (acc=0.7701, ‘1-hamming’=0.9502, LRAP=0.9541) are higher than for the contextualised model (acc=0.7407, ‘1-hamming’=0.9410, LRAP=0.9485). This means that the behaviours displayed by a texter or volunteer in one message do not predict the behaviour of the other actor in the subsequent message.

**Table 5:** Results of behaviour classification for individual messages. Comparison of two models: without context and with message context, i.e., including the text of the preceding and following messages.

| Model | Accuracy (weighted) | 1-hamming loss | LRAP |
| --- | --- | --- | --- |
| no context | **0.7701** | **0.9502** | **0.9541** |
| $\pm 1$ message | 0.7407 | 0.9410 | 0.9485 |

This result is in contrast with Experiment 1, where we found that context improved the prediction of conversation stages, since conversation stages are likely to remain unchanged for several messages. On the other hand, behaviours are unique to each message and rapidly varying across time and between actors, hence our metrics suggest that identifying behaviours based only on the current message is sufficient and appropriate. While there may be relationships between behaviours from message to message not currently picked up by our model, it is understandable that textual information from the volunteer (in a preceding or subsequent message) may not inform classification of texter behaviour, or vice-versa, in the current message. The behaviours of two consecutive messages are unlikely to be the same (and even less likely when including three messages), due to the interchange of actors across text messages and the rapid variation of behaviours.

The high accuracy demonstrated for classification of texter and Volunteer behaviours suggests that the codebook of defined categories of behaviour displayed by texters and volunteers are meaningful and distinct. The ability to classify psychologically-relevant behaviours at scale is the first step for future work to determine the relationship between such behaviours and important features such as conversation outcomes or different texter profiles.

### 3.3 Revealing texter demographics using full conversation classification (Experiment 3)

Finally, we examine the classification of entire conversations using the texter survey results as ground truths (see Section 2.2.3). In particular, we focus on particular texter demographics of special interest to better understand the users of the Shout service.

#### 3.3.1 Model optimisation and prediction performance

To build and test a capable model for conversation level classification, we chose three texter demographic variables of interest: (1) Age: 13 or under/over 13; (2) Autism: self-identification of an autism diagnosis or not, (3) Non-binary gender: self-identification of gender as non-binary, versus all other answers to the question on gender. These choices of demographic subgroups was motivated by their interest to MHI experts.

The conversation-level classification models for the different demographic survey questions were fine-tuned at the checkpoint of the MLM. The classification accuracy of the three trained models for each binary label are reported on validation data (Table 6) with excellent performance (above 95% accuracy). Revealingly, while the results of texter surveys showed that 6.63%, 6.18%, and 4.94%, of texters were, respectively, of age 13 or under, autistic, and non-binary gender, the model-predicted proportions for the whole data set were 4.35%, 1.28%, and 1.60%, respectively. This suggests that younger texters, and/or those who have been diagnosed with autism and/or identify as having non-binary gender may be more likely than others to complete the survey.

**Table 6:** Classification performance for three demographic variables. We report the proportion of texters that self-identified in each demographic category in the survey. Using the trained models, we then predicted the class of the remaining (unlabelled) conversations and report the percentage of total texters predicted in each demographic category. We also report the difference between the survey-reported and predicted proportions of each category.

| Demographic category | Accuracy (weighted) | Survey Proportion | Predicted Proportion | $\Delta$ Proportion |
| --- | --- | --- | --- | --- |
| Age 13 or under | 0.9575 | 0.0663 | 0.0435 | -0.0228 |
| Autism | 0.9533 | 0.0618 | 0.0128 | -0.0490 |
| Non binary gender | 0.9592 | 0.0494 | 0.0160 | -0.0334 |

#### 3.3.2 Feature analysis and interpretation

We used LIME to provide insight into the classification models for each demographic class. For the age label, LIME indicates that, as could be expected, numbers less than or equal to 13 are predictive of being aged 13 or less, and likely occur in response to the volunteer asking the texter’s age, which can occur when the volunteer suspects they are talking to a child, for safeguarding purposes. Additionally, words that refer to school life, such as ‘school work’, ‘bullying’ or ‘lesson’ also appear as important, and may indicate that bullying and school pressures are key issues faced distinctly by young texters. Moreover, we find that words corresponding to friends and parents are predictive of the texter age (Figure 5(a)).

**Figure 5:**
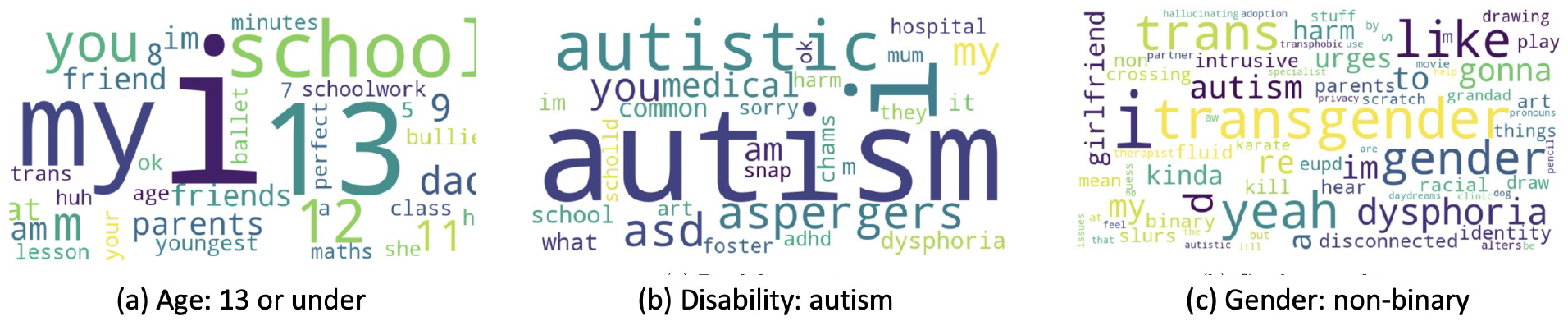
Word cloud for words that best explains the model predictions for (a) age group 13 or under, (b) autism and (c) non-binary gender.

Application of LIME to the autism classification model identified obvious words (Figure 5(b)), such as ‘autistic’, ‘autism’ and ‘aspergers’, but words representing other disabilities that are not directly related to autism were also present in the word cloud, such as ‘adhd’ or ‘dysphoria’. This may suggest that texters with these disabilities tend to express themselves in similar ways, or that texters with autism are more likely to have other diagnoses, which is a well-known phenomenon [46].

For the non-binary gender class label, (Figure 5(c)), we again find obvious words as predictive, including ‘transgender’ and ‘trans’; however, less obvious words such as ‘gonna’, ‘yeah’ and ‘like’ appear as important. We also see that ‘autism’ is quite a significant word in the word cloud of non-binary gender. This implies that individuals with non-binary genders are more likely to discuss autism in their conversations with Shout compared to cisgender individuals, which is consistent with the co-occurrence of autism and gender dysphoria previously documented [47, 48, 49].

## 4 Discussion

We have demonstrated the use of NLP to gain insights into users and service provision of a digital mental health crisis service. We trained and tested our NLP models using a unique large-scale data set from the Shout crisis text messaging service. To our knowledge, this is the first demonstration of NLP to generate predictions of attributes of mental health crisis conversations at both a message-level and conversation-level, including conversation stages, psychologically-relevant behaviours and texter demographics.

While increasing numbers of people are turning to non-traditional forms of mental health support, few services are evaluated for impact or quality. In our first experiment, we developed a model to predict the ‘conversation stage’ of each text message. Shout Volunteers are trained to follow six distinct conversation stages to guide a texter from a place of distress to a calmer place, and assessing adherence to this structure is a potential indicator of general conversation quality and volunteer skill. Our model is able to accurately predict (weighted accuracy = 87.75%) the conversation stage of messages on an unseen test set, which, given the randomised sampling of annotated conversations, should extrapolate to unannotated conversations. Analysing the structure of conversations has previously been shown as predictive of conversation outcome. Althoff *et al*.[7] derived linguistic features related to conversation outcomes that allowed the identification of optimal conversation strategies for volunteers. Although our analysis of conversation stages has some similarities to this work, we do not rely on feature engineering and instead derive a latent space embedding of conversations using a deep learning NLP model. Further, our analysis of conversation structure examines both Volunteer and texter, whilst Althoff et al. focussed only on the volunteer[7].

As well as a potential marker of conversation quality or efficacy, the importance of accurate prediction of conversation stages opens several other opportunities. For example, coaches could use the predicted stages of conversations as a tool to review conversations with volunteers and improve their training. Moreover, using the predicted conversation stage for each message, one could potentially track conversation stages in real time and direct prompts to volunteers or their Clinical Supervisors if the conversation appears ‘stuck’, or stages appear to be missed. Additionally, the conversation structures and timings can be compared for different issues or subgroups (e.g., in conversations with extremely high levels of distress or suicidal ideation), or for conversations rated as helpful or unhelpful by texters. Such comparisons could generate insights into conversation formats that are most appropriate for texter needs, allowing for appropriate guidance for Volunteers to tailor conversations, and for supporting those in mental health crisis more broadly.

The Shout data set also provides potential for insight into the psychologically-relevant behaviours of people in mental distress as expressed during a conversation, and behaviours that are most helpful from a supporter. Language-based deficits are common symptoms of mental health crises and associated behaviours[50], and NLP techniques can generate accurate numeric embeddings of human-developed behavioral codes (i.e. annotated behavioural categories) [23]. The ability to label at scale the psychologically-relevant behaviours of Shout texters and volunteers would enable deeper exploration of behaviours demonstrated by different texter groups (e.g., those experiencing different levels of distress) and the volunteer behaviours most predictive of helpful outcomes. Here we demonstrate the successful development of an NLP model to predict the presence of up to three of six psychologically-relevant behaviours (setting intention, enquiring, expressing distress, reflecting, corresponding, discord) in each message from a volunteer or texter. The model performs with 95.02% prediction accuracy. Such a result supports the validity of the behavioural classifications developed using a qualitative inductive thematic analysis[32, 33] and their distinctiveness, and shows they can be predicted from linguistic features. Our results complement a previous study examining behaviours of texters using data from the USA Crisis Text Line [51]. Using machine-learning techniques the researchers queried message-level data to identify conversations that included phrases related to suicidality and revealed three distinct behaviours among texters expressing suicide risk. Here, we present a more general study with a wider range of behaviours and included all texters and volunteers within the analysis, not just those at high risk. The ability to predict behaviours of both texters and Volunteers will allow in future work for examination of texter-volunteer interactions and their relative efficacy (relationship to conversation outcomes), and whether volunteers do and/or should adapt their behaviours to different texters.

It is worth noting that while message-level predictions could potentially be made in real time, the current technique of context concatenation sets a limit on predicting messages, i.e., a lag of specifically one time step (message). A possible solution for achieving real-time monitoring is to fine-tune a unidirectional NLP model such as GPT-3 [12], which is pre-trained on an auto-regressive language modelling task so that only previous context is considered to predict the current word.

Finally, we also considered prediction at a conversation level. The Shout post-conversation survey collects demographic data on perceived conversation helpfulness scores from texters, but only a minority of texters complete the survey, and there is considerable opportunity for survey bias[52, 53, 54]. Our results show that NLP models can also predict with high accuracy three different conversation survey results (age 13 or under: 95.75%; autism: 95.33%; non-binary gender: 95.92%), based on the conversation content as a whole. This allows us to determine demographic data for the entire texter cohort. The high accuracy for each demographic class, all with imbalanced classes, suggests the model should achieve similarly high accuracy on other survey results, and could support development of novel triaging strategies for crisis services, or help with the stratification of different population subgroups for further in-depth analysis of their experiences. Whilst we focussed on predicting conversation demographics here, our analysis could be extended to other features such as suicide risk or conversation helpfulness in future work[17, 18, 7].

Using the predicted population demographics, we observe that the true demographic breakdown of Shout users differed from the survey results—the predicted proportions were lower across all tested demographic categories, suggesting survey bias for some demographic categories. Texters identified as autistic or with non-binary gender were over-represented 5 × in the survey relative to the predicted results. As it is important to understand who is using digital mental health services and experiencing mental health crisis, the ability to generate predicted texter demographics without survey bias provides vital information to inform service development. Further, the age at which younger individuals develop mental health problems and expressions of mental health crisis in children is still not well understood[55, 56]. Identifying the complete set of conversations made by young texters could offer an invaluable data set to evaluate mental health crises in younger texters, complementing machine learning studies aimed at predicting mental health issues in children and adolescents[57, 58].

While we have provided initial results in the use of NLP to analyse and monitor mental health conversations, there are potential limitations to our study. First, we had limited annotations for message-level data (both conversation stage and behavioural keys, but not for conversation-level demographic data) due to the manual labelling of messages being resource intensive for the clinically trained research psychologist. Hence, increasing the number of training samples would improve model performance. Additionally, although several Shout clinical professionals were involved in the iterative development of a codebook for annotation of psychologically-relevant behaviours that produced consistent annotations on a conversation subset (validation), our final message-level annotations were performed by a single clinical research psychologist which could potentially lead to bias in the train and test data[59]. Despite this, the feature importance analysis returned words that sensibly reflected what would be expected for each class, suggesting that the model learned class categories did not appear to contain bias. Having provided a proof of concept for NLP, a key future aim will be to annotate more conversations and to do so with multiple raters.

Looking forward, we aim to train models to classify labels that could be considered directly actionable or provide a direct measure of conversation efficacy. For example, a key future aim is prediction of a texter’s risk of suicide, Furthermore, we would like to predict the extent to which a conversation de-escalated a texter or mitigated their crisis, which would help us measure the efficacy of the Shout service. Moreover, using the predicted conversation stages, we aim to use Bayesian modelling[60] to understand how conversation structure may confer conversation outcomes, and use unsupervised learning[61] to identify natural clusters of conversations that might not relate to ground truth labels.

Overall, our study highlights the potential for NLP to help gain insights into mental health service provision and the experiences of people in mental and emotional crisis. The ability to accurately predict message level attributes of crisis text conversations provides novel insights into psychologically-relevant behaviours displayed by individuals in mental 7distress and those supporting them, and the structures of those conversations. The demonstrated prediction of post-conversation survey results from conversation content has wide implications for monitoring, triaging and mental health service improvement, as well as for understanding the complex interactions of texter demographics. Taken together, this application of NLP to a unique large-scale charity mental health data set opens routes for monitoring and improvement of digital mental health services, and to gain novel insights, at scale, for those who most need our help.

## Data Availability

Data used in the present study is not available due to its sensitive nature.

## 5 Acknowledgments

We thank Ovidiu Serban, Daniel Cahn, David Newman, Joy Li, Jianxiong Sun, Jingru Yang and Sophia Yaliraki for valuable discussions. We thank Mental Health Innovations for providing training and access to their invaluable data set on their highly secure server environment. We thank Lia Maragou and Lauren Duncan for their help in codebook development for data annotation.

## 7 Author Contributions

ZL, RP and EL conceived the study. AN produced the message-level annotations. ZL conducted all machine learning and natural language processing analyses, supervised by RP, EL, AN, MU and MB. ZL, EL, RP and MB drafted the manuscript and all authors reviewed the manuscript before submission.

